# Routine antimicrobial therapy for severe fever with thrombocytopenia syndrome does not improve prognosis: overlap weighting analysis using a nationwide inpatient database

**DOI:** 10.1101/2024.06.14.24308904

**Authors:** Satoshi Kutsuna, Hiroyuki Ohbe, Hiroki Matsui, Hideo Yasunaga

## Abstract

**Background:** Severe fever with thrombocytopenia syndrome (SFTS) is an infectious disease that presents a formidable challenge due to the absence of established therapeutic strategies that are explicitly tailored to its management. This study aimed to assess the impact of routine antimicrobial therapy on patients diagnosed with SFTS in Japan. We conducted a comprehensive retrospective cohort analysis using extensive data from a national inpatient database.

**Methodology/Principal Findings:** This study scrutinized data from adult patients with SFTS and categorized them based on whether they received antimicrobial treatment within the initial 2 days of hospital admission. A meticulous evaluation was carried out on a range of outcomes, such as in-hospital mortality rates, overall costs associated with hospitalization, and length of hospital stay. Overlap weighting was applied along with multivariate regression models to enhance the reliability of the findings through confounder adjustment. The outcomes showed no significant improvement in the prognosis of patients with SFTS who received routine antimicrobial therapy. The use of antimicrobials did not yield statistically significant improvements in in-hospital mortality rates or other secondary outcomes, suggesting that such therapeutic interventions may not be necessary during the early stages of hospital admission.

**Conclusion/Significance:** This study underscores that the indiscriminate administration of antimicrobials does not substantively enhance the prognosis of patients with severe SFTS, hence dissuading the universal implementation of antimicrobials as a conventional component of therapeutic strategies in managing severe cases of SFTS.

**Author summary:** Severe fever with thrombocytopenia syndrome is a tick-borne infection that is endemic in East Asia, including Japan, China, and South Korea. At present, there are no effective antiviral drugs against SFTS. It has been reported that patients who received antimicrobial agents for SFTS early on had a better prognosis. To validate this study, we compared SFTS patients who received antimicrobials within 2 days of admission with those who did not, using Japanese DPC data. Overlap weighting was applied along with multivariate regression models to enhance the reliability of the findings through confounder adjustment. The results showed no difference in in-hospital mortality, hospitalization costs, or hospital days between the two groups. In conclusion, the analysis of patients with SFTS using a nationwide inpatient database revealed that routine antimicrobials did not improve patient prognosis. Further, our findings do not support the universal use of antimicrobials in patients with severe SFTS.

## Introduction

Severe fever with thrombocytopenia syndrome (SFTS) is an infectious disease characterized by a high mortality rate. SFTS is caused by the SFTS virus, a novel phlebovirus identified as a prominent public health concern [1]. This disease presents a formidable challenge due to the absence of established therapeutic strategies that are explicitly tailored to its management. Whether favipiravir improves the prognosis of SFTS has not yet been established [2], and the benefits of steroid pulse therapy are also unknown [3].

A notable characteristic of SFTS is the substantial reduction in white blood cells, which has been linked to heightened susceptibility to co-infections, such as secondary bacterial infections [4]. These complications further exacerbate the clinical course of the disease, making the management of patients with SFTS particularly challenging. Recent international studies have suggested that early administration of antibacterial agents can substantially improve the prognosis of patients with SFTS [5]. If validated, this notion could mark a pivotal shift in the approach to manage this daunting infection. In light of these considerations, our study aims to examine this proposition. Using the Japanese Diagnosis Procedure Combination (DPC) database, we conducted a comprehensive analysis to ascertain whether the early administration of antibacterial agents influences the survival outcomes of patients with SFTS.

## Methods

### Data source

This retrospective cohort study was conducted using data from the DPC database, a nationwide inpatient database in Japan. This database contains discharge abstracts and administrative claims data voluntarily contributed by over 1,200 acute care hospitals in Japan [6]. In addition, it contains all inpatient-level data, including age, sex, diagnosis recorded using the International Classification of Diseases, Tenth Revision (ICD-10) codes, daily procedures recorded using Japanese medical procedure codes, daily drug administration, admission status, and discharge abstracts. Previous validation studies of this database have shown high specificity and moderate sensitivity for diagnoses and high specificity and sensitivity for procedures [7].

### Study population

Using the Japanese DPC inpatient database from July 2010 to March 2021, we identified adult patients diagnosed with SFTS according to the ICD-10 code A938. Patients with suspected SFTS were excluded. Patients younger than 20 years of age and those discharged within 2 days of admission were excluded.

### Treatment

Patients who received antimicrobials within 2 days of admission were categorized as the antibiotic group. Patients who did not receive antimicrobials within 2 days of admission were categorized as the control group.

### Outcomes and covariates

The primary outcome was in-hospital mortality rate. The secondary outcomes were the total hospitalization cost (1 US dollar equivalent to 115 Japanese yen) and length of hospital stay.

The covariates included age, sex, smoking history, body mass index (BMI) at admission, physical function at admission measured using the Barthel Index (BI) score [8], Japan Coma Scale score at admission [9], Charlson Comorbidity Index score [10], fiscal year at admission, ambulance use, admission on a weekend (i.e., on Saturday or Sunday) or Japanese holiday, diagnosis of hemophagocytic syndrome at admission (ICD10 codes D762 or D763), diagnosis of sepsis at admission, and organ support therapies within 2 days of admission (including intensive care unit/high-dependency care unit admission, supplemental oxygen, mechanical ventilation, catecholamine, renal replacement therapy, red blood cell transfusion, fresh frozen plasma transfusion, and platelet transfusion). BMI was categorized as <18.5, 18.5–24.9, 25.0–29.9, and ≥30.0 kg/m^2^, or missing data. Physical function at admission was categorized as total/severe dependence (BI: 0–60), slight/moderate dependence (BI: 61–99), or independent (BI = 100). The Japan Coma Scale correlated well with the Glasgow Coma Scale, and the scores were categorized as alert consciousness, confusion, somnolence, or coma [9]. The Charlson Comorbidity Index was scored based on the diagnosis for each patient, and the scores were categorized as 0, 1, 2, or ≥3 [10].

### Statistical analysis

Overlap weighting was our primary approach for comparing the outcomes between the antibiotic and the control groups [11,12]. Overlap weighting is a recently developed propensity scoring method that attempts to mimic the important attributes of randomized clinical trials and addresses some of the issues of the classic inverse probability of treatment weighting. Overlap weighting emphasizes the target population with the greatest overlap in the observed characteristics between the two groups.

A multivariate logistic regression model using all the abovementioned covariates was employed to compute the propensity scores for patients who received antimicrobials within 2 days of admission. Patients in the antibiotic and control groups were weighted by the probability of not receiving (propensity score, 1) and receiving (propensity score) treatment, respectively. To assess the performance of the overlap weighting, all the covariates were compared using standardized differences, with absolute standardized differences ≤10% considered to denote negligible imbalances between the two groups [13]. Risk differences and 95% confidence intervals of the outcomes were calculated using weighted generalized linear models, irrespective of the outcome type. For 30-day in-hospital mortality, we generated Kaplan–Meier curves in the weighted cohort and performed a log-rank test of equality.

### Sensitivity analysis

We performed sensitivity analysis by changing the definition of the antibiotic group. Only patients who received cefepime, piperacillin/tazobactam, and carbapenem were included in the antibiotic group, and those who did not receive antimicrobials within 2 days of admission were included in the control group.

All analyses were performed using the Stata/MP 17.0 software (StataCorp). Continuous variables are presented as medians and interquartile ranges, and categorical variables are presented as numbers and percentages. All reported *P* values were two-sided, and *P* < 0.05 was considered statistically significant.

### Ethics approval and consent to participate

This study was approved by the Institutional Review Board of the University of Tokyo (approval number: 3501-3; December 25, 2017). No information permitting the identification of patients, hospitals, or physicians was obtained, and the requirement for informed consent was waived because of the anonymous nature of the data.

## Results

We identified 451 patients hospitalized due to SFTS from 149 hospitals during the study period. After excluding 6 patients aged <20 years and 20 patients who were discharged within 2 days of admission, a total of 425 eligible patients were included. Of these, 287 (68%) who received antimicrobials within 2 days of admission were allocated to the antibiotic group, and the remaining 138 (32%) were allocated to the control group. In the antibiotic group, the patients received a median of 4 days (interquartile range, 2–6 days) of antimicrobial treatment. In the control group, 42 (30%) patients received antimicrobial treatment after day 3 of admission.

The baseline characteristics of the patients before and after overlap weighting are presented in **Table 1**. Before overlap weighting, patients in the antibiotic group were more likely to be older, obese, have higher physical function at admission, have worse Japan Coma Scale scores at admission, have sepsis at admission, and receive organ support therapies compared to those in the control group. The distributions of propensity scores before and after overlap weighting are shown in **Figs S1 and S2**. After overlap weighting, the patient characteristics were perfectly balanced between the two groups (**Table 1**).

**Table 1.**
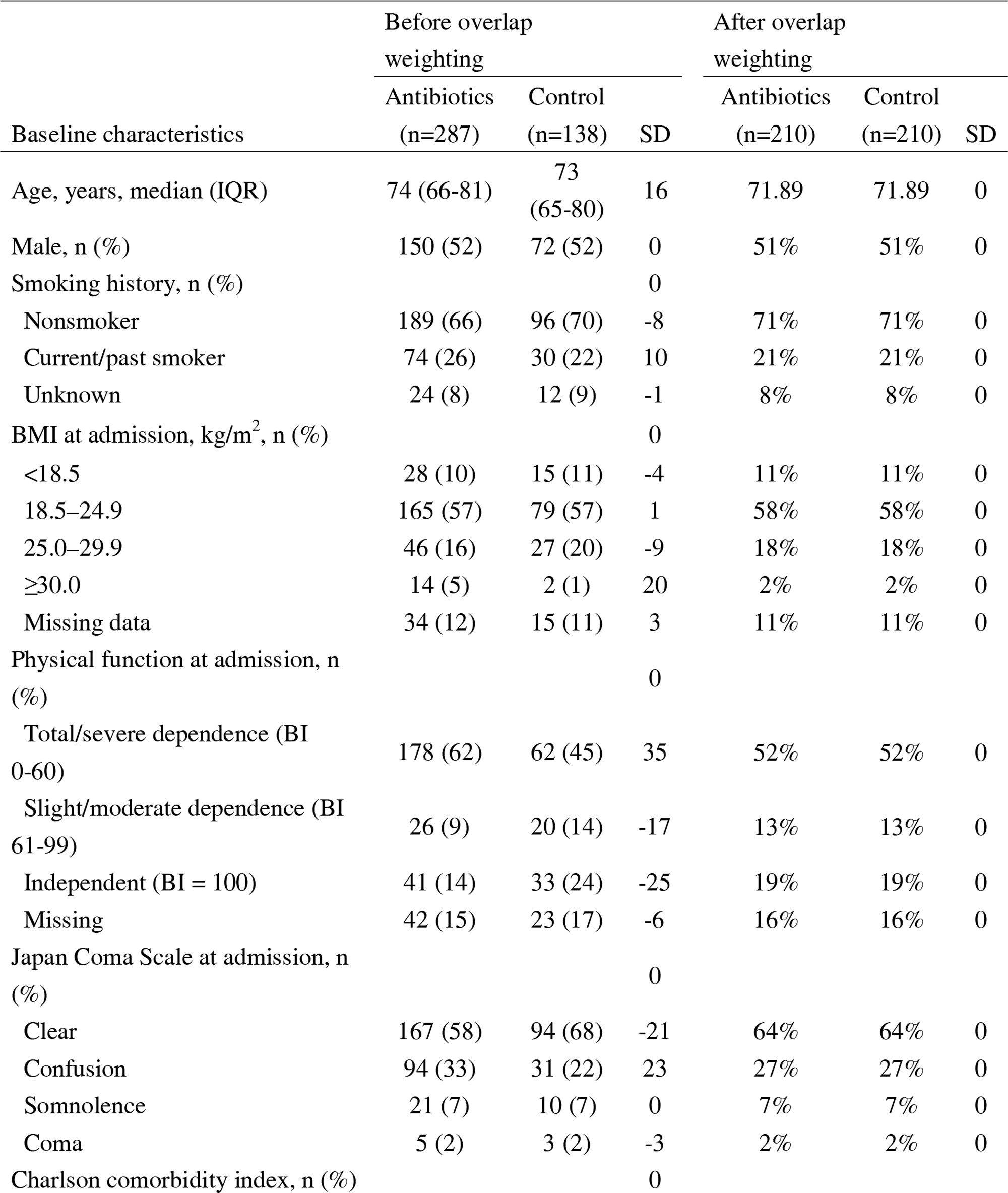
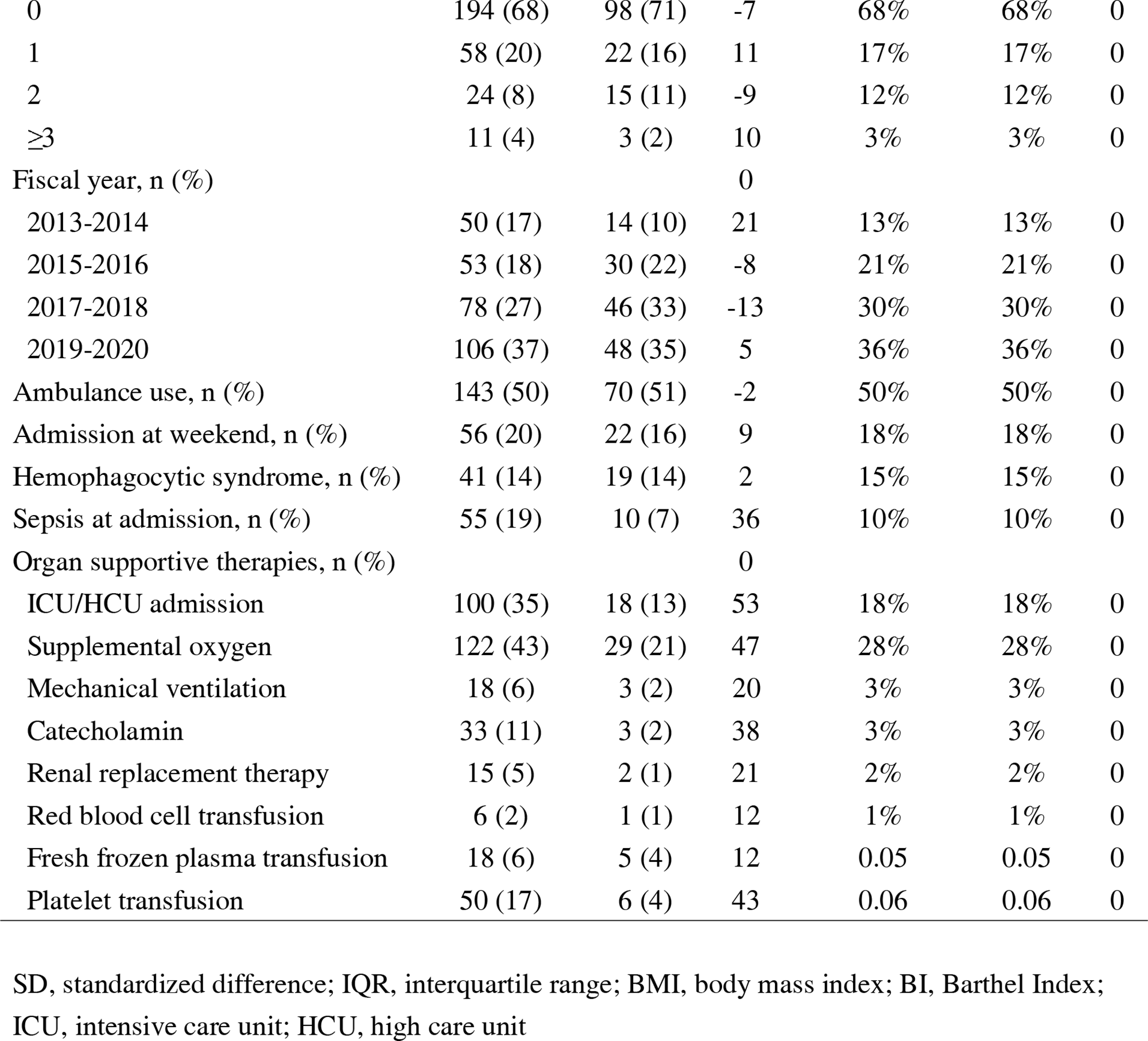
Patient characteristics before and after overlap weighting.

| Baseline characteristics | Before overlap weighting |  |  | After overlap weighting |  |  |
| --- | --- | --- | --- | --- | --- | --- |
|  | Antibiotics<br>(n=287) | Control<br>(n=138) | SD | Antibiotics<br>(n=210) | Control<br>(n=210) | SD |
| Age, years, median (IQR) | 74 (66-81) | 73<br>(65-80) | 16 | 71.89 | 71.89 | 0 |
| Male, n (%) | 150 (52) | 72 (52) | 0 | 51% | 51% | 0 |
| Smoking history, n (%) |  |  | 0 |  |  |  |
| Nonsmoker | 189 (66) | 96 (70) | -8 | 71% | 71% | 0 |
| Current/past smoker | 74 (26) | 30 (22) | 10 | 21% | 21% | 0 |
| Unknown | 24 (8) | 12 (9) | -1 | 8% | 8% | 0 |
| BMI at admission, kg/m<sup>2</sup>, n (%) |  |  | 0 |  |  |  |
| <18.5 | 28 (10) | 15 (11) | -4 | 11% | 11% | 0 |
| 18.5–24.9 | 165 (57) | 79 (57) | 1 | 58% | 58% | 0 |
| 25.0–29.9 | 46 (16) | 27 (20) | -9 | 18% | 18% | 0 |
| ≥30.0 | 14 (5) | 2 (1) | 20 | 2% | 2% | 0 |
| Missing data | 34 (12) | 15 (11) | 3 | 11% | 11% | 0 |
| Physical function at admission, n (%) |  |  | 0 |  |  |  |
| Total/severe dependence (BI 0-60) | 178 (62) | 62 (45) | 35 | 52% | 52% | 0 |
| Slight/moderate dependence (BI 61-99) | 26 (9) | 20 (14) | -17 | 13% | 13% | 0 |
| Independent (BI = 100) | 41 (14) | 33 (24) | -25 | 19% | 19% | 0 |
| Missing | 42 (15) | 23 (17) | -6 | 16% | 16% | 0 |
| Japan Coma Scale at admission, n (%) |  |  | 0 |  |  |  |
| Clear | 167 (58) | 94 (68) | -21 | 64% | 64% | 0 |
| Confusion | 94 (33) | 31 (22) | 23 | 27% | 27% | 0 |
| Somnolence | 21 (7) | 10 (7) | 0 | 7% | 7% | 0 |
| Coma | 5 (2) | 3 (2) | -3 | 2% | 2% | 0 |
| Charlson comorbidity index, n (%) |  |  | 0 |  |  |  |
| 0 | 194 (68) | 98 (71) | -7 | 68% | 68% | 0 |
| 1 | 58 (20) | 22 (16) | 11 | 17% | 17% | 0 |
| 2 | 24 (8) | 15 (11) | -9 | 12% | 12% | 0 |
| ≥3 | 11 (4) | 3 (2) | 10 | 3% | 3% | 0 |
| Fiscal year, n (%) |  |  | 0 |  |  |  |
| 2013-2014 | 50 (17) | 14 (10) | 21 | 13% | 13% | 0 |
| 2015-2016 | 53 (18) | 30 (22) | -8 | 21% | 21% | 0 |
| 2017-2018 | 78 (27) | 46 (33) | -13 | 30% | 30% | 0 |
| 2019-2020 | 106 (37) | 48 (35) | 5 | 36% | 36% | 0 |
| Ambulance use, n (%) | 143 (50) | 70 (51) | -2 | 50% | 50% | 0 |
| Admission at weekend, n (%) | 56 (20) | 22 (16) | 9 | 18% | 18% | 0 |
| Hemophagocytic syndrome, n (%) | 41 (14) | 19 (14) | 2 | 15% | 15% | 0 |
| Sepsis at admission, n (%) | 55 (19) | 10 (7) | 36 | 10% | 10% | 0 |
| Organ supportive therapies, n (%) |  |  | 0 |  |  |  |
| ICU/HCU admission | 100 (35) | 18 (13) | 53 | 18% | 18% | 0 |
| Supplemental oxygen | 122 (43) | 29 (21) | 47 | 28% | 28% | 0 |
| Mechanical ventilation | 18 (6) | 3 (2) | 20 | 3% | 3% | 0 |
| Catecholamin | 33 (11) | 3 (2) | 38 | 3% | 3% | 0 |
| Renal replacement therapy | 15 (5) | 2 (1) | 21 | 2% | 2% | 0 |
| Red blood cell transfusion | 6 (2) | 1 (1) | 12 | 1% | 1% | 0 |
| Fresh frozen plasma transfusion | 18 (6) | 5 (4) | 12 | 0.05 | 0.05 | 0 |
| Platelet transfusion | 50 (17) | 6 (4) | 43 | 0.06 | 0.06 | 0 |

SD, standardized difference; IQR, interquartile range; BMI, body mass index; BI, Barthel Index; ICU, intensive care unit; HCU, high care unit

The outcomes before and after the overlap weighting are shown in **Table 2**. After overlap weighting, no statistically significant difference in in-hospital mortality was observed between patients in the antibiotic and control groups (17.4% vs. 14.3%; risk difference: +3.1%; 95% confidence interval: −3.9% to +10.1%). The Kaplan–Meier curve comparing the two groups showed a higher mortality rate in the antimicrobial group, although the difference was not significant (log-rank test, *P* = 0.409) (**Fig 1**). No statistically significant differences between the groups in total hospitalization costs (6826 USD vs. 6239 USD; risk difference: 810 USD; 95% confidence interval: −1615 USD to +7818 USD), and length of hospital stay (13 days vs. 15 days; risk difference: 1.2 days; 95% confidence interval: −3.8 days to +6.1 days) were observed.

**Table 2.** Outcomes before and after overlap weighting.

|  | Before overlap weighting | After overlap |
| --- | --- | --- |

|  |  |  | weighting |  |  | P<br>value |
| --- | --- | --- | --- | --- | --- | --- |
|  | Antibiotics<br>(n=287) | Control<br>(n=138) | Antibiotics<br>(n=210) | Control<br>(n=210) | Difference<br>(95% CI) |  |
| In-hospital mortality, n (%) | 62 (21.6) | 18 (13.0) | 37 (17.4) | 30 (14.3) | 3.1% (-3.9, 10.1%) | 0.38 |
| Total hospitalization cost, USD | 8107<br>(4839-16017) | 5947<br>(3298-9844) | 6829<br>(4104, 11693) | 6239<br>(3527, 10462) | 810<br>(-1615, 7818) | 0.513 |
| Length of hospital stay, days | 14 (7-23) | 15 (8-21) | 13 (8-23) | 15 (8-22) | 1.2 (-3.8, 6.1) | 0.647 |

Of 287 patients who received antimicrobials within 2 days of admission, 109 patients received cefepime, piperacillin/tazobactam, and carbapenem. The sensitivity analysis results after changing the definition of the antibiotic group were similar to those of the main analyses (**Table 3**).

**Table 3.**
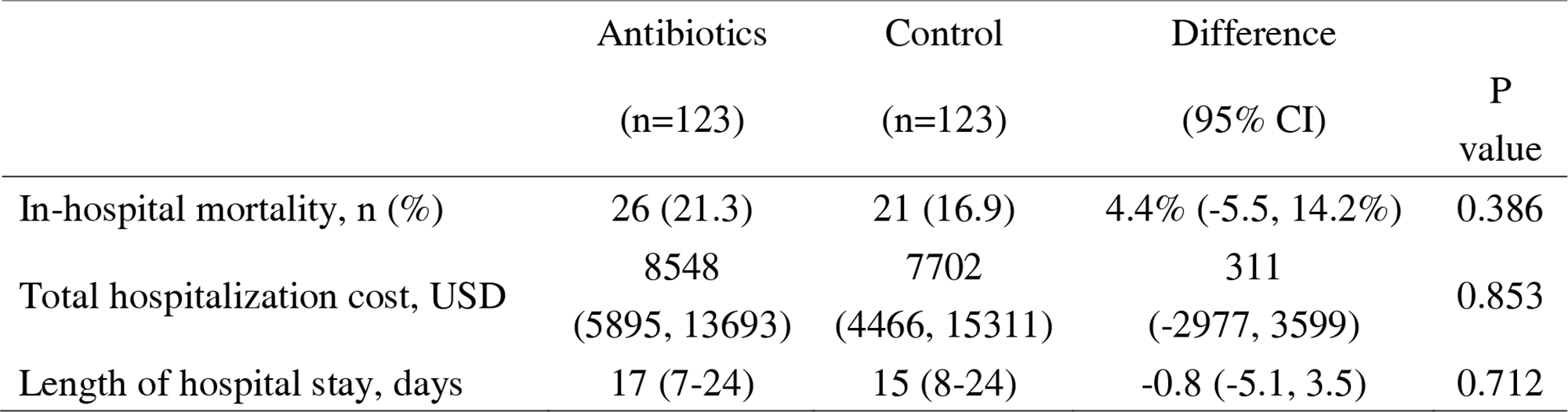
Outcomes of the sensitivity analysis.

## Discussion

In our study, routine antimicrobial therapy for SFTS did not improve the prognosis. Similarly, no differences were found in the sensitivity analysis that focused on the antimicrobial agents cefepime, piperacillin/tazobactam, and carbapenem.

Xia et al. showed that in 377 patients, neurological symptoms and joint index scores were the two indicators associated with mortality [5]. Further, they found that patients with SFTS who received antimicrobials had a lower mortality rate than those who did not. Even in patients without marked granulocytopenia or infection, the mortality rate in the subgroup meeting only one of the two mortality-related indicators was only 7.2% in the group treated with antimicrobials and 47.4% in the control group (*P* < 0.001), and early prophylaxis was associated with lower mortality (9.0% vs. 22.8%, *P* = 0.008). These findings contradict the results of the present study. Notably, the sample size in the study by Xia et al. was limited and they did not adjust for severity by covariates and conducted simple comparisons. SFTS is an infectious disease characterized by a decrease in the white blood cell count in the acute phase. As fever is also a frequent symptom in SFTS, patients with SFTS often fit in the febrile neutropenia criteria of absolute neutrophil count of <1000/μL [14]. SFTS is often associated with bacterial complications [4,15,16]. Therefore, clinicians may be tempted to administer antimicrobials prophylactically to patients with SFTS. Notably, 68% of patients with SFTS enrolled in the study received antimicrobials within 2 days of admission. However, routine administration of antimicrobial agents did not improve the prognosis. Furthermore, no differences were observed when the analysis was limited to antimicrobial agents such as cefepime, piperacillin/tazobactam, and carbapenem, which are recommended for febrile neutropenia. Our analysis indicates routine administration of antimicrobials should not be recommended to patients with SFTS from the time of admission, regardless of the white blood cell count.

Increasing antimicrobial resistance due to the inappropriate use of antimicrobial agents is a pressing concern. Surprisingly, 68% of patients with SFTS received antimicrobial therapy within day 2 of hospitalization. Antimicrobial therapy is often unavoidable when Japanese spotted fever and tsutsugamushi disease are not ruled out, as they are often prevalent simultaneously in SFTS-prevalent places [17]. In Japan, diagnosis of Japanese spotted fever and tsutsugamushi disease requires a testing request from the regional health laboratory center, which often takes time to obtain test results and inevitably requires longer periods of antimicrobial administration. To minimize the administration of unnecessary antimicrobial agents as much as possible, establishing rapid testing methods for Japanese spotted fever and tsutsugamushi disease is essential. Our study had some limitations. First, because blood test results were not available for our study using DPC data, the present analysis was not limited to SFTS patients with decreased white blood cell counts. We defined the antibiotic group as cases in which antimicrobials were initiated within 2 days of admission. The antibiotic group included patients who received antimicrobials even though their leukopenia was not marked. The control group included patients who received antimicrobials after leukopenia became pronounced on day 3 of hospitalization or later. Therefore, whether empirically administered antimicrobials that are initiated after leukopenia becomes pronounced following hospitalization are effective remains unclear. Second, SFTS is occasionally complicated by secondary fungal infections, particularly invasive pulmonary aspergillosis [18,19]. We also attempted to analyze the relationship between the use of antifungal agents within 2 days of hospitalization and prognosis but could not continue due to the small number of patients involved (n = 25). Third, we used a multicenter, real-world database in Japan. Consequently, the initiation of antimicrobials was contingent on the decision of the attending physician, potentially leading to confounding by indication. We controlled for measured confounders using overlap weighting; however, unmeasured confounders may still persist. The severity of the disease was not comprehensively adjusted, possibly leading to a skewed higher mortality rate in the antibiotic group owing to the inclusion of more severe cases. Fourth, despite including cases categorized as SFTS in the database, we cannot ascertain whether the diagnoses were made using PCR. The gold standard for diagnosing SFTS in Japan is the PCR test, which is performed at regional health laboratories. Considering that SFTS diagnoses are typically not made without laboratory testing, it is commonly presumed that disease registration in the Japanese Diagnosis Procedure Combination Database relies on PCR test results. Despite these limitations, the strengths of our study include a larger sample size, and we also adjusted for severity using covariates.

In conclusion, the analysis of patients with SFTS using a nationwide inpatient database revealed that routine antimicrobials did not improve patient prognosis. Further, our findings do not support the universal use of antimicrobials in patients with severe SFTS.

## Data Availability

All data produced in the present study are available upon reasonable request to the authors

## Authorship statement

All the authors met the ICMJE authorship criteria. SK and HO drafted the manuscript HO was the chief investigator responsible for the data analysis. All authors contributed to the writing of the final manuscript. All authors critically revised the manuscript, commented on drafts of the manuscript, and approved the final report.

---

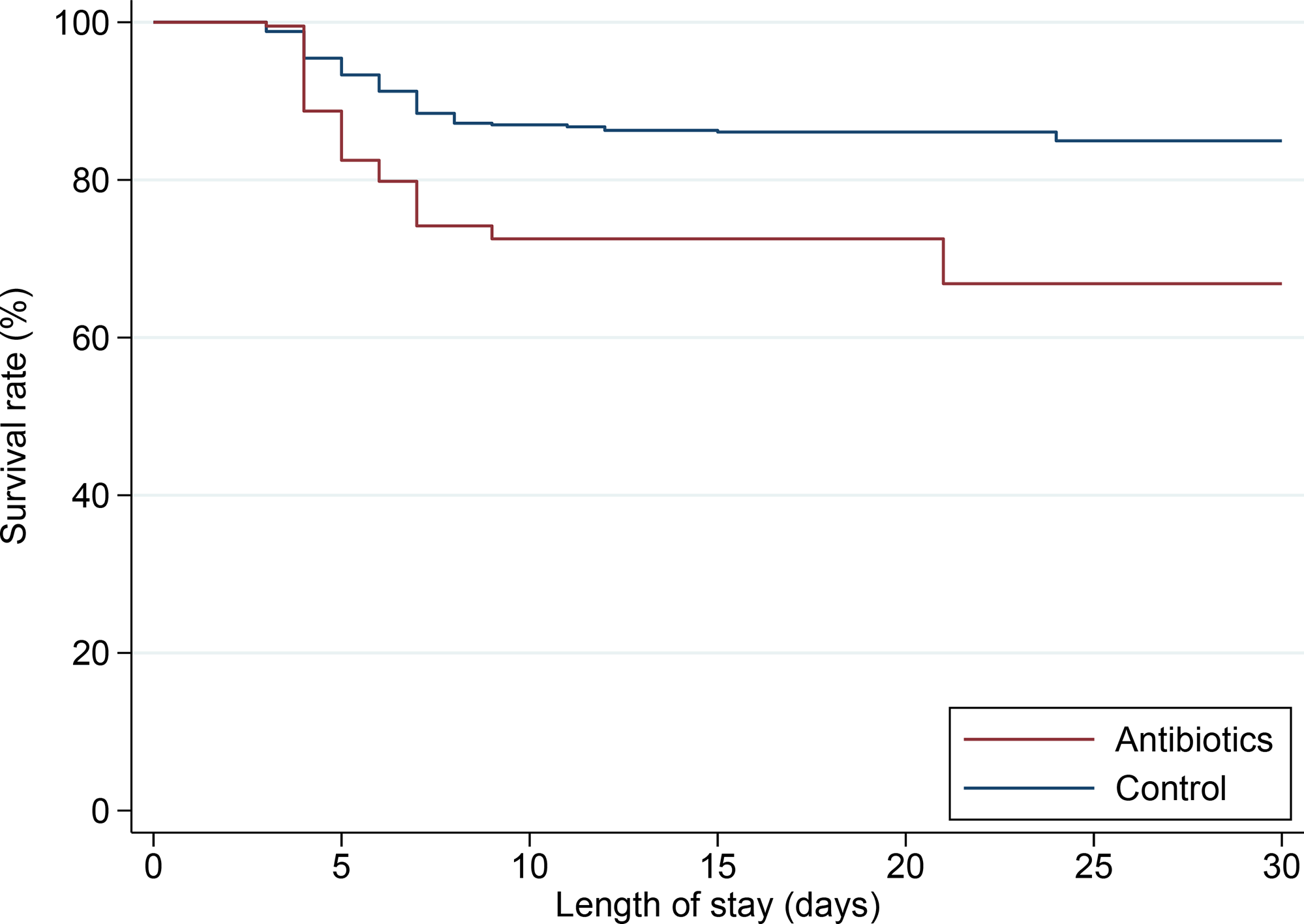

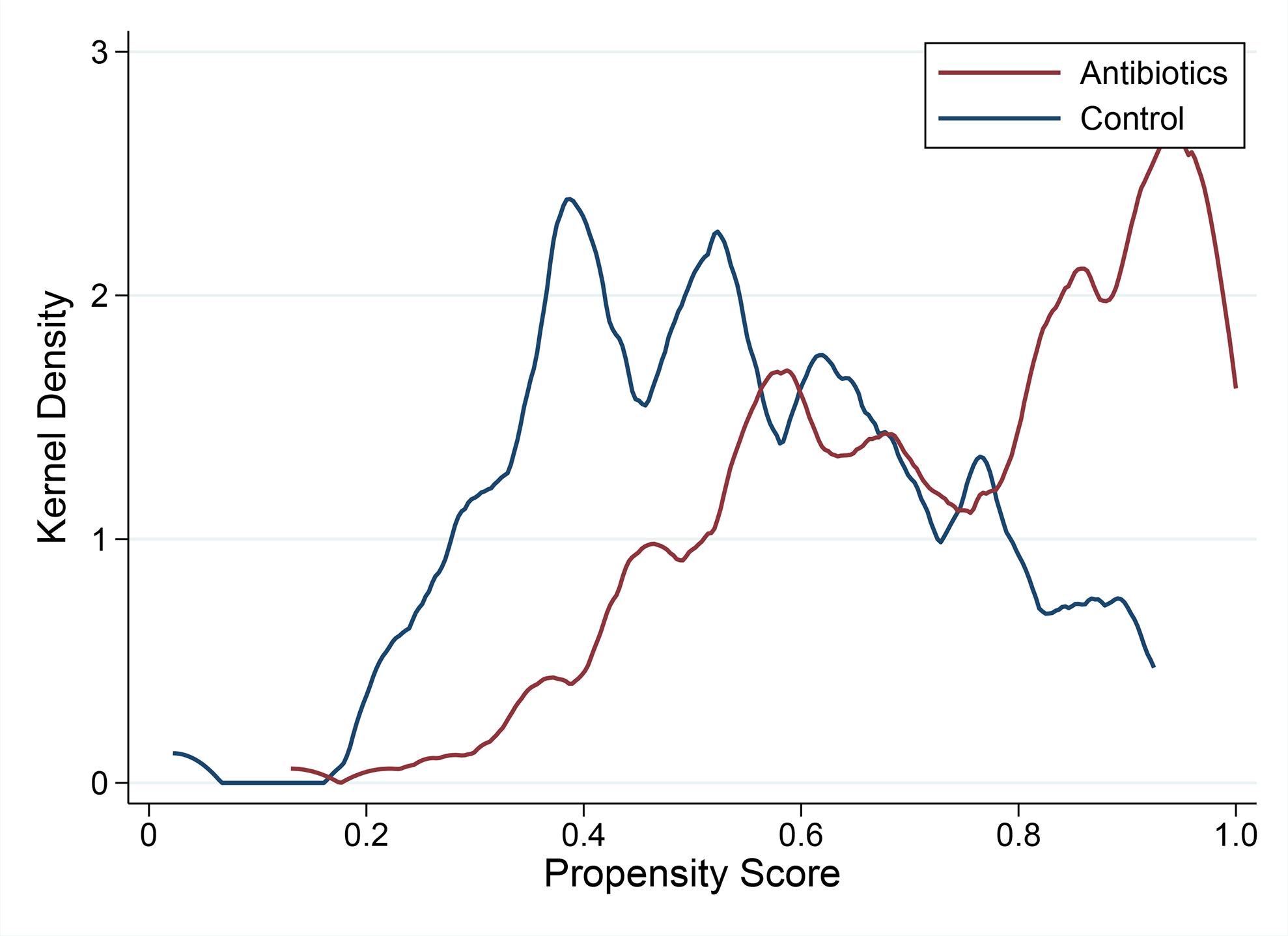

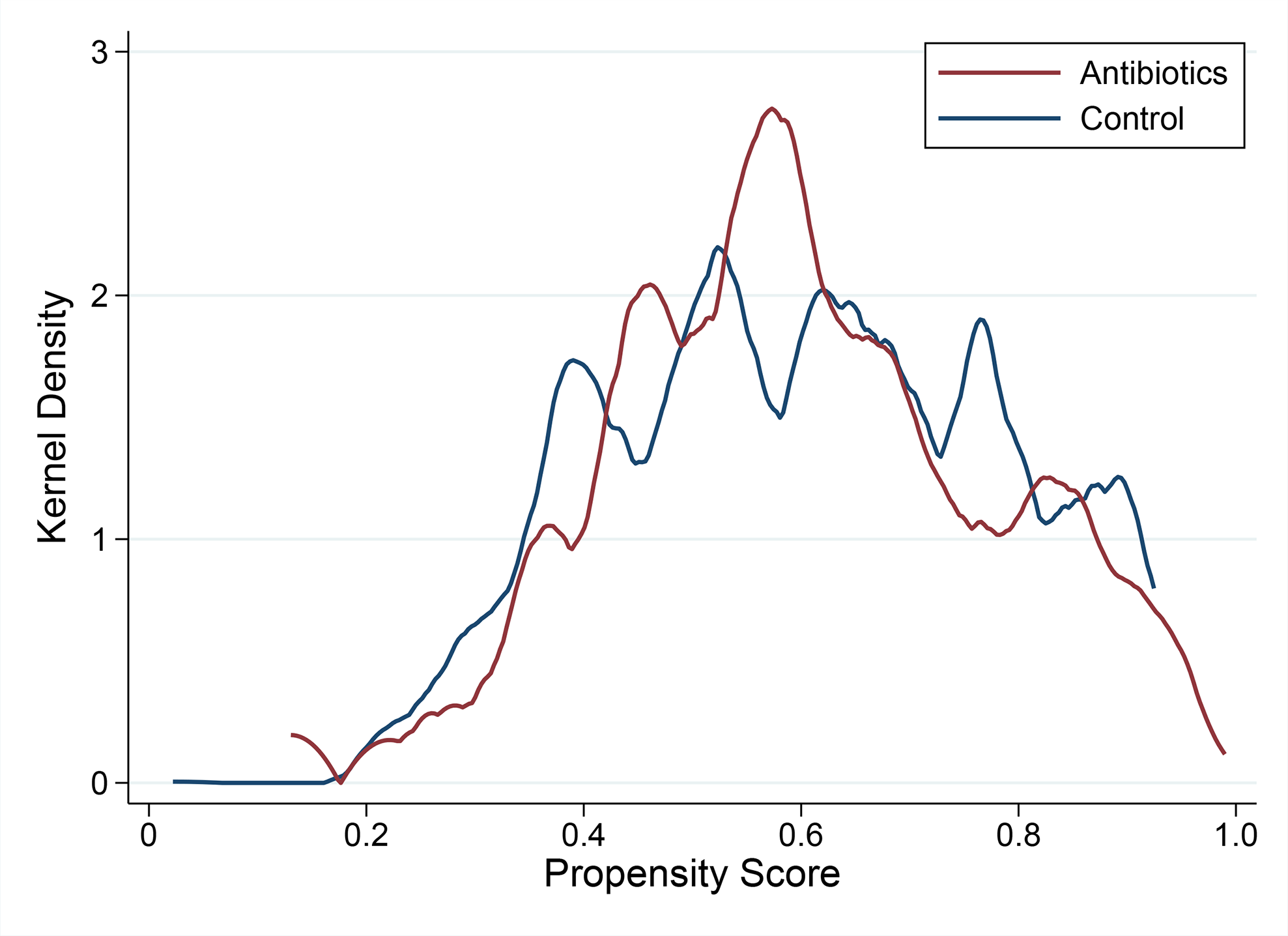

